# Standardizing Phenotypic Algorithms for the Classification of Degenerative Rotator Cuff Tear from Electronic Health Record Systems

**DOI:** 10.1101/2024.09.29.24314565

**Authors:** S Herzberg, NE Garduno-Rapp, H Ong, S Gangreddi, A Chandrashekar, W Wei, L LeClere, W Wen, K Hartmann, N Jain, A Giri

## Abstract

**Background:** Degenerative rotator cuff tears (DCTs) are the leading cause of shoulder pain, affecting 30% to 50% of individuals over 50. Current phenotyping strategies for DCT rely on heterogeneous combinations of procedural and diagnostic codes, leading to concerns about misclassification. We aimed to create universal phenotypic algorithms to classify DCT status across EHR systems.

**Methods:** Using Vanderbilt University Medical Center’s de-identified EHR system, we developed and validated two sets of algorithms—one requiring imaging verification and one without imaging verification—to identify DCT cases and controls. The algorithms used combinations of ICD and CPT codes, as well as natural language processing (NLP), to increase diagnostic certainty. Manual chart reviews by trained personnel blinded to case-control determinations were conducted to compute positive (PPV) and negative predictive values (NPV).

**Results:** The algorithm development process resulted in five phenotypes with an overall predictive value of 94.5%. By approach: 1) code only cases that required imaging confirmation (PPV = 89%), 2) code only cases that did not require imaging verification (PPV = 92%), 3) NLP-based cases that did not require imaging verification (PPV = 89%), 4) code-based controls that were confirmed by imaging (NPV = 90%), and 5) code and NLP-based controls that did not require imaging verification (NPV = 100%). External validation demonstrated 94% sensitivity and 75% specificity for the code-only algorithms. The addition of NLP increased the number of identified cases without compromising predictive values.

**Conclusions:** These algorithms represent an improvement over current phenotyping strategies and allows for EHR studies of unprecedented size

## INTRODUCTION

Rotator Cuff Disease (RCD) or Rotator Cuff Syndrome (RCS), is a composite term often used to describe multiple related pathologies of the rotator cuff, including subacromial pain (impingement) syndrome, rotator cuff tendinopathy, and symptomatic partial and full thickness rotator cuff tears.[1] It is among the most common causes of pain and disability.[2] However, the heterogeneity of the term RCD is a major limitation of rotator cuff research as it includes a variety of conditions as a composite outcome, therefore associating risk factors uniformly across all included conditions despite meaningfully different pathophysiologic mechanisms of each injury. Moreover, cuff tears range in presentation from debilitating pain to asymptomatic incidental findings with some studies reporting rates of asymptomatic tears that account for up to 65% of all tears, making it difficult to assess tear status.[3,4]

Importantly, current studies investigating rotator cuff tear rely either on hospital-based case control/cohort studies[5–15], which are time consuming, expensive and result in small sample sizes, or single diagnostic codes [16–19] for the classification of rotator cuff tears and as a result suffer from high risk of bias. Moreover studies using procedural and diagnostic codes suffer from three main limitations: 1) Phenotypic heterogeneity across studies as outlined above. 2) Lack of validated algorithms specifically for degenerative rotator cuff tears (DCT) that can be used broadly across institutions utilizing electronic health records (EHRs), so GWAS conducted to date do not use consistent definitions.[10,11,16,18] 3) Current phenotyping strategies have involved use of single ICD-9/10 and Read v2/v3 codes. These codes are recognized to be imprecise and often assigned to patients without definitive musculoskeletal diagnoses,[20] a problem also highlighted in work from our group.[21] This previous work identifies key features from EHRs that are predictive of cuff tear status individually, however, falls short of constructing clearly defined algorithms (specific combinations of structured or unstructured codes) that can be applied and tested to classify cases and controls for broader use in EHRs across the US. To date, only one EHR-based study, conducted by Yanik et al.[22], has limited their phenotypic definition to include only procedural codes specific to degenerative cuff repair and employed temporal limitations to ensure exclusion of traumatic tears. However, this study did not perform manual validation of their algorithm. To our knowledge, no validated, published algorithm can be used as standardized methods to classify DCT across different EHR systems. This is particularly important as adoption of EHRs for high-throughput research is on the rise. Our investigation builds on previous work to develop and validate algorithms for application in EHRs allowing varying levels of query. Table 1 summarizes the main phenotypic definition categories currently used in studies evaluating genetic predisposition to rotator cuff tears and provides a non-comprehensive list of studies in each category.

**Table 1.**
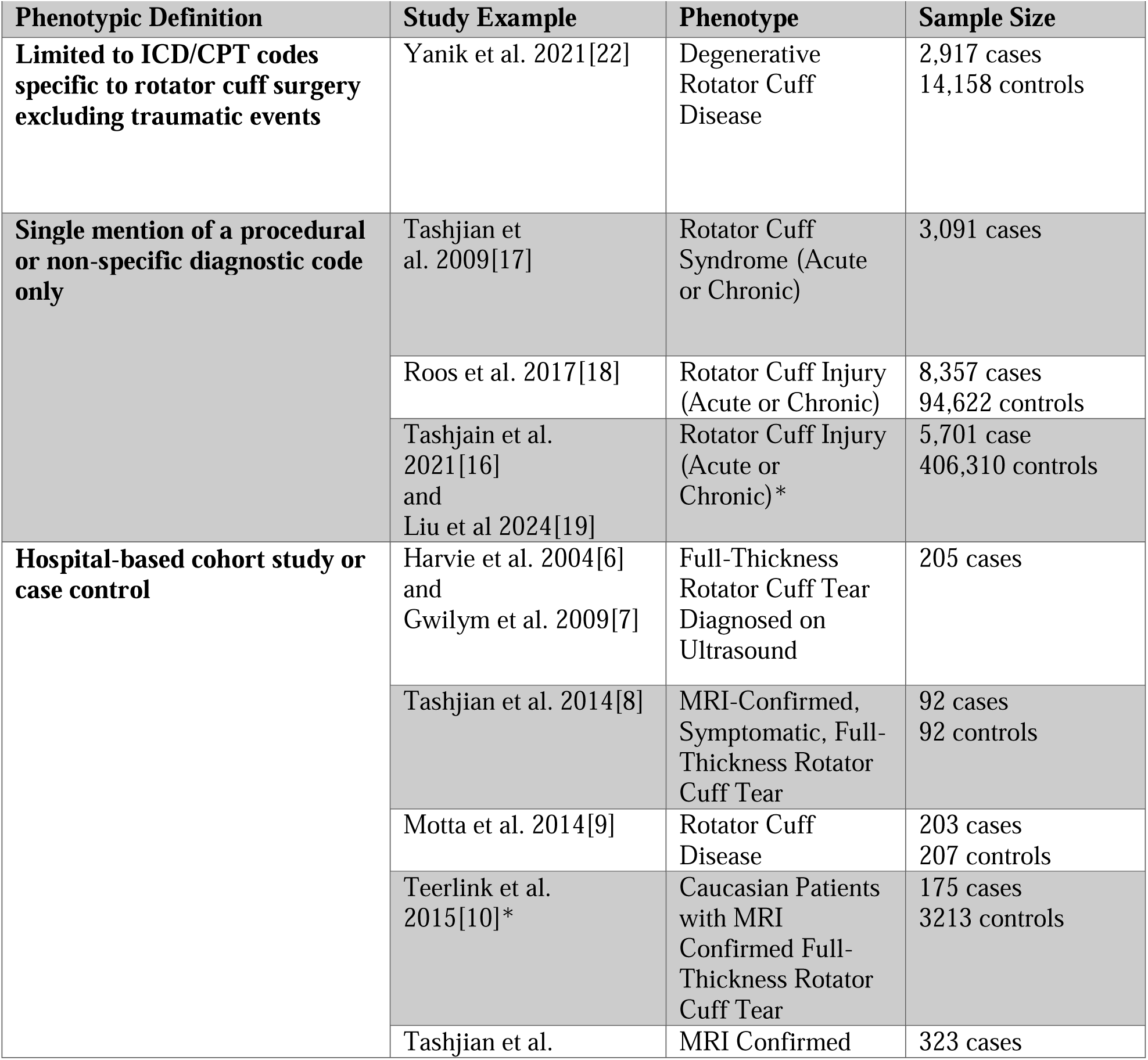
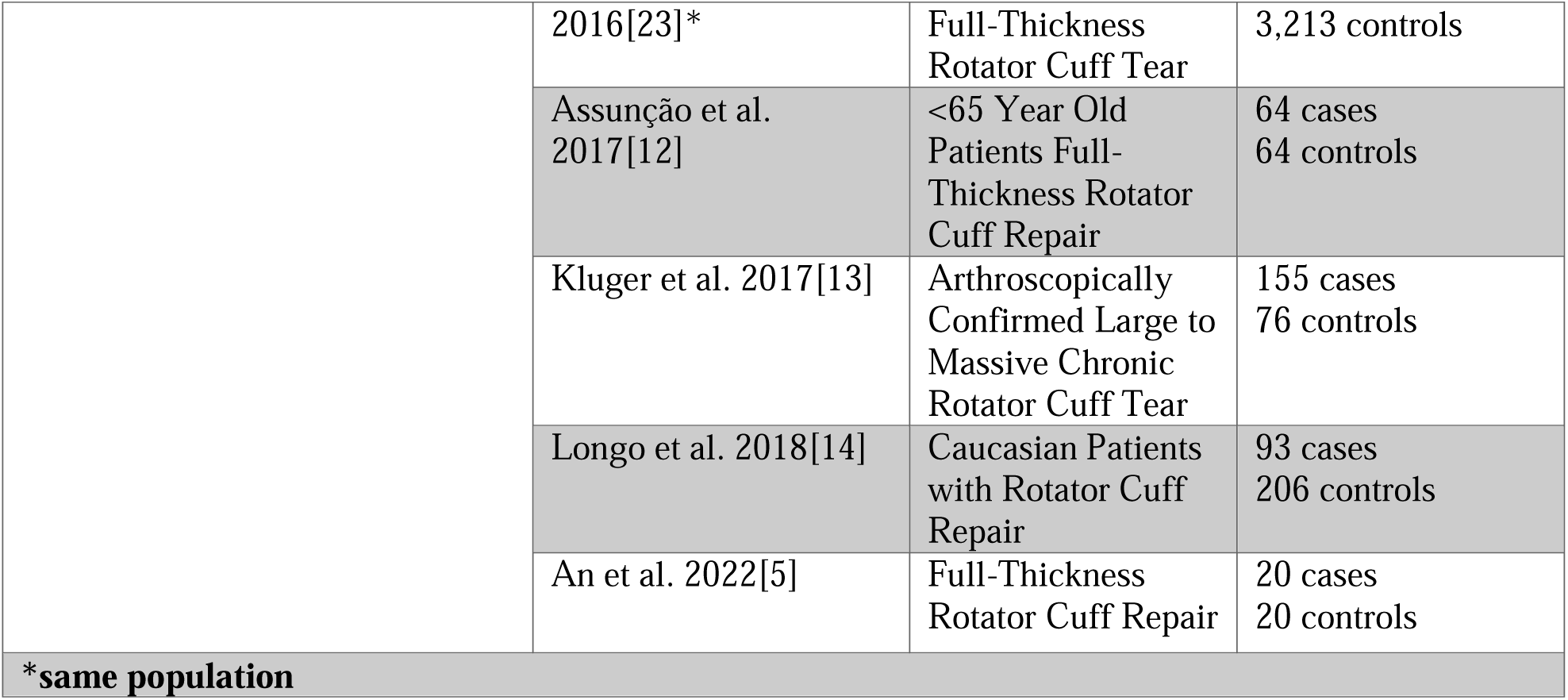
Phenotypic Assessment of Currently Published Studies on Genetics of Rotator Cuff Tear.

Digitization of electronic health records and linkage with biorepositories, coupled with widespread advances in biomedical informatics, now allow for unprecedented access to large-scale health-care data. Leveraging EHR systems and linked biorepositories along with principled phenotyping approaches can provide larger sample sizes and can allow inquisition of research questions that were previously intractable.[24–26] As recently demonstrated by our team for diabetic retinopathy, this approach has the potential to advance clinical and etiological understanding of health conditions.[27] However, to date, there are no validated phenotypic algorithms designed to classify rotator cuff tear status using EHRs and current phenotypic selection strategies for rotator cuff tears rely on a heterogeneous combination of billing and diagnostic codes[16–19,22] which may identify composite outcome variables for rotator cuff disease rather than tears, leading to concern for misclassification of cases and controls. In addition to heterogeneity due to outcome definition, substantial variability in data availability constraints across EHR systems (availability of notes, access to images, access to billing codes only etc.) further prevent transferability and reproducibility of algorithms. The purpose of this study is to provide principled approaches to conduct high quality, reproducible research on rotator cuff tears through the development of validated EHR algorithms for identifying DCT cases and controls.

## METHODS

We developed and validated multiple algorithms to identify cases of DCT and controls utilizing the Vanderbilt University Medical Center de-identified EHR database, the Synthetic Derivative (SD) to accommodate applicability to different EHR systems that have varying degrees of constraints in data availability. We used the SD, a de-identified database derived from Vanderbilt’s electronic medical records, containing clinical data for over 2.2 million individuals[26], to develop and validate two broad types of algorithms that optimize the classification of DCT—one type requiring evidence of image verification (as documented by the presence of tear on MRI, CT-arthrogram, or ultrasound of the shoulder in EHR record) referred to here on out as ‘imaging-confirmed’, and others that did not require chart documented image verification, referred to as ‘imaging-not-required’ (Figure 1). Recognizing varying limits in access, and computational capability, we designed the non-imaging-based algorithms with the option of utilizing natural language processing (NLP) to perform mining of charts, referred here on out as ‘NLP-based’. Any algorithms which do not utilize NLP and is entirely reliant on diagnostic and procedural codes such as ICD-9, ICD-10 and CPT codes will be referred to as ‘code-based’ algorithms. Among the algorithms that do not require imaging, we additionally evaluated the impact of incorporating natural language processing (NLP) on sample size and predictive values (Figure 1).

**Figure 1.**
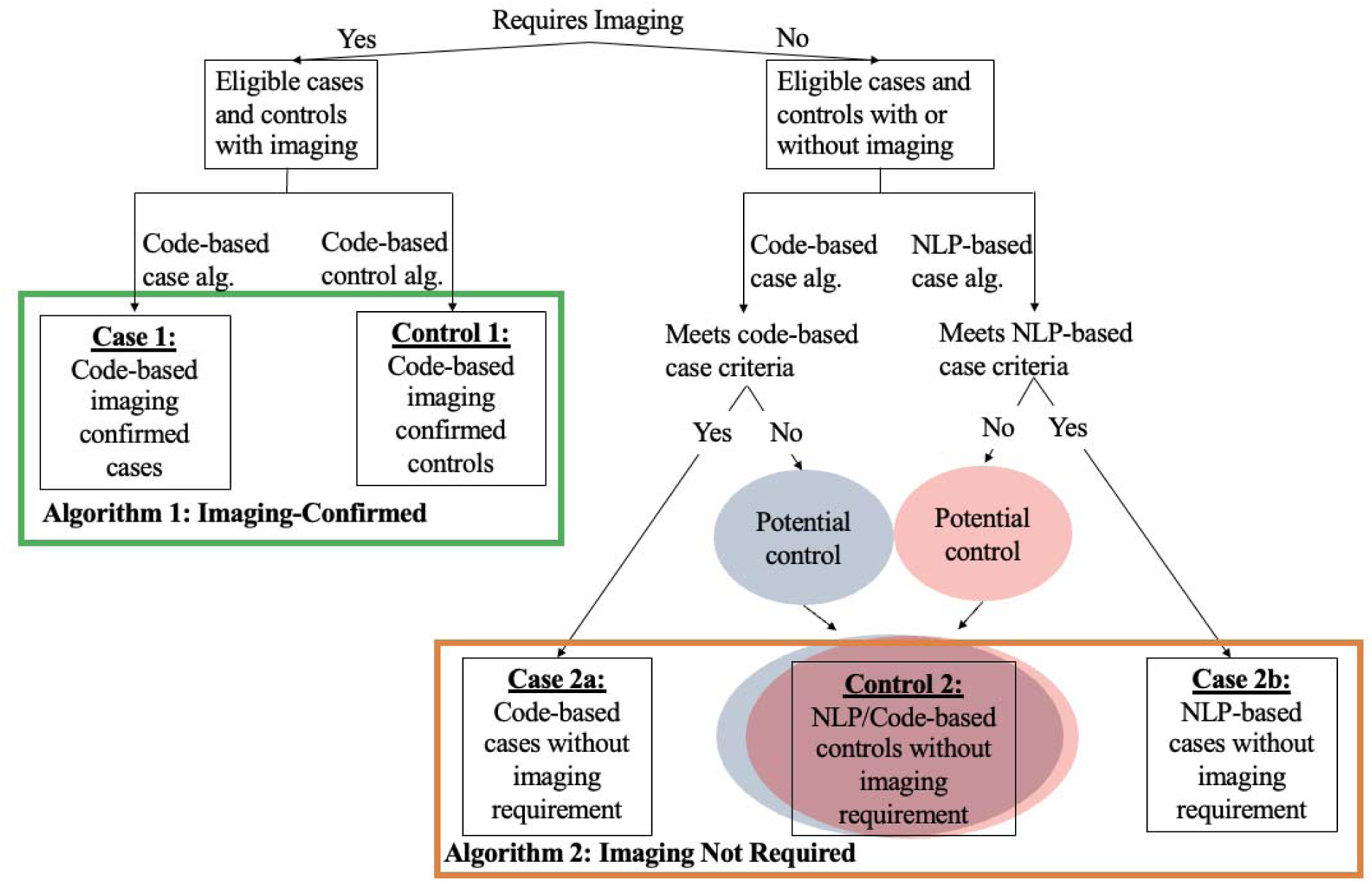
Flow Chart for Classification of Cases and Controls by Algorithm.

### Development

The algorithm development process, which took place in the VUMC SD, included application of preliminary algorithms, several rounds of manual review, performance testing, algorithm refining and re-evaluation, and then final validation. A flowchart summarizing the process for algorithm development and validation is shown in Figure 2. This was an iterative development process that resulted in several rounds of assessment and over 4,500 case and control charts reviewed before achieving final algorithm status and conducting formal validations

**Figure 2.**
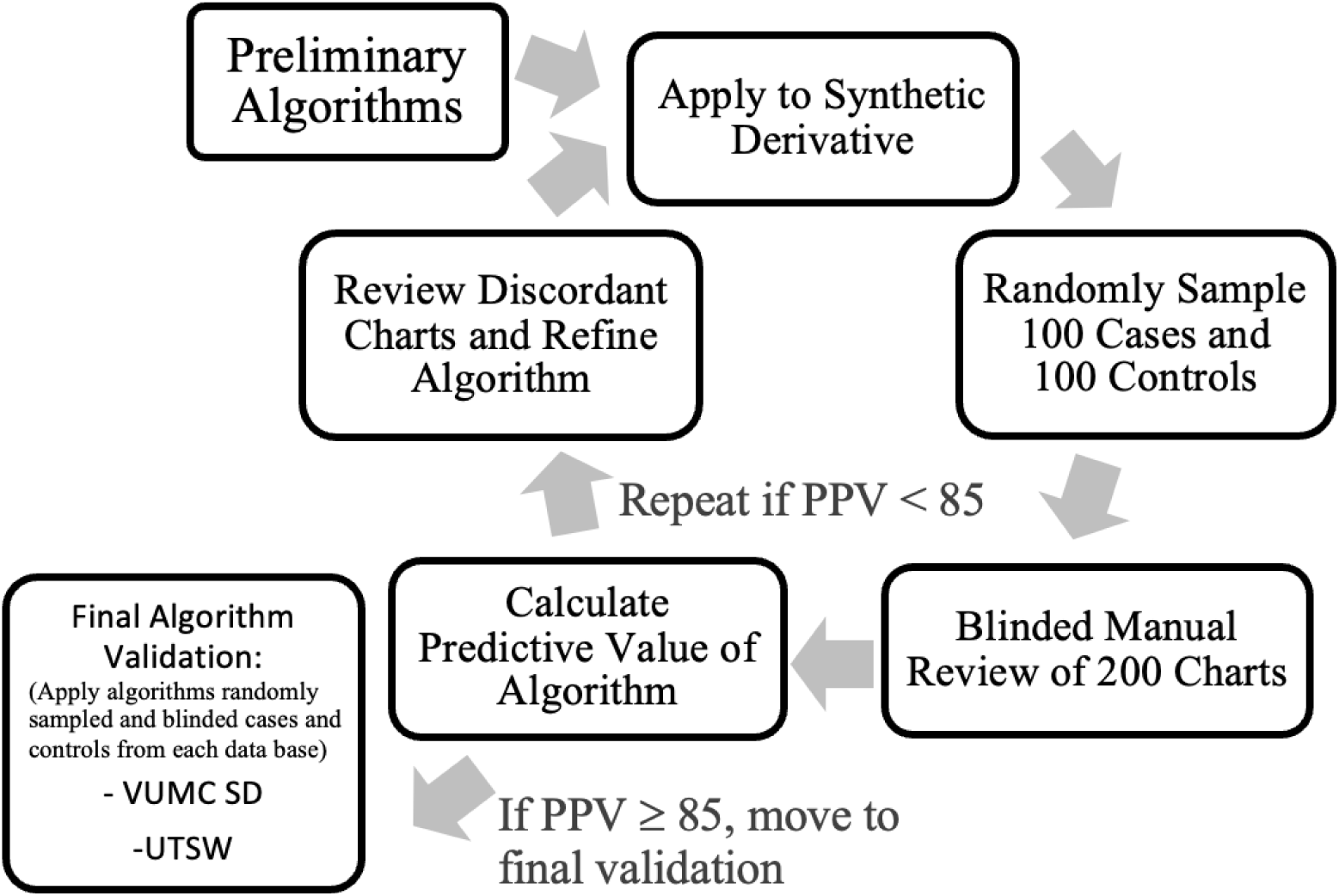
Algorithm Development.

Informed by previous work,[21] clinical experience, and *a priori* knowledge of medical record documentation of rotator cuff diseases, each phenotype consisted of an initial set of sub-criteria designed to capture (for cases) or exclude (for controls) a substantial variety of ways in which rotator cuff tears could have been documented in the EHR system. Each of these initial phenotypes underwent five full rounds of iterative manual review for validation before final formatting for formal blinded dual validation. Each round of review consisted of a random selection of 100 cases and 100 controls from each different case and control phenotype. Over the course of an iterative review, a total of 4,500 charts were reviewed. Throughout the iterative review, the phenotypes were amended to optimize performance, and any sub-criteria with less than 85% predictive value were either modified and re-validated or dropped from analysis before being formalized for final review. Results of the review process and final phenotype structure are outlined below for each of the five phenotypes.

### Validation

Upon satisfactory development of primary algorithms (at least 85% predictive value during iterative review), the resulting final algorithms, presented in the results, were internally validated by applying the algorithm to the entire SD database and then to randomly selected 100 algorithmically determined cases and controls from each sub-criteria. This process resulted in a final validation set of 700 charts, 400 cases (100 NLP-based imaging-not-required cases, 200 code-based imaging-not-required cases, and 100 code-based imaging confirmed cases) and 300 controls (100 code-based imaging confirmed controls, and 200 code and NLP-based imaging-not-required controls). 200 code-based imaging-not-required cases and 200 code and NLP-based imaging-not-required controls were selected in order to allow sufficient sizes to assess the predictive value of each sub-criteria. A stratified random sampling strategy was selected to ensure the sufficient number of cases and controls were selected from each sub-category to allow for a more precise estimation of predictive values for both cases and controls.

### Manual Review Criteria

Randomly selected case and control samples were combined, and manual chart review was performed to determine true case and control status by two trained reviewers who were blinded to algorithm-based determinations of case-control status. Reviewers, who had access to the patient’s de-identified electronic medical record, read through all provider notes, imaging reports, surgical procedural notes and outside medical record notes/communications, to determine true case-control status. Patients were included as a case if they had a diagnosis of a rotator cuff tear as documented by mention of cuff tear in an imaging report, indication of prior rotator cuff repair or surgical note indicating rotator cuff repair, or a provider-documented confirmation of rotator cuff tear diagnosis. Patients were excluded as cases if the cuff tear was documented as acute, or if there was indication of a major traumatic shoulder event directly preceding the tear diagnosis without indication of antecedent cuff pathology. Patients were included as controls if there was no evidence of a rotator cuff tear anywhere in their electronic medical record. Patients were included as image-confirmed controls if they had an imaging report that documented an intact rotator cuff, or if they had a provider note that indicated they underwent an imaging or surgical procedure and were noted to have an intact cuff.

After dual review, any discordant charts were resolved by an orthopedic surgeon who served as the expert third party reviewer. Resultant case versus control assessment was then used to calculate the final Positive Predictive Value (PPV = true positives/ (true positives+ false positives)), Negative Predictive Value (NPV= true negatives/ (true negatives + false negatives)) as well as the C-statistic (C= true positives/ (1-false positives)) [28].

### External Validation

External validation was conducted using participants from the CuffGEN study recruited at the University of Texas Southwestern Medical Center (UTSW). Because the NLP ecosystem was not yet implemented at UTSW EHR system, only the code-based (imaging-confirmed) algorithms could be applied for validation. Briefly, CuffGEN is an ongoing multi-center NIH-funded study recruiting patients with imaging-verified rotator cuff tear cases and controls across 4 medical centers across the US: Vanderbilt University Medical Center, UTSW, Massachusetts General Brigham, and University of Michigan. At the time of external validation, the USTW portion of the CuffGEN study consisted of 492 participants (405 cases of rotator cuff tears and 87 controls). Cases were over the age of 40 and had evidence of cuff tear on MRI that was atraumatic in nature based on chart documented history of the disease. Controls consisted of individuals who presented for evaluation of shoulder pathology that were over the age of 40 with MRI verified lack of rotator cuff tear. Notably, controls could have had a condition other than a cuff tear, such as adhesive capsulitis, osteoarthritis, or shoulder instability. Because the algorithm was applied to the CuffGEN cohort, with a case-control status determined *apriori*, sensitivity and specificity were used to evaluate the performance of the algorithm.

## ETHICAL CONSIDERATIONS

This study was approved by the Institutional Review Board (IRB #140857). To protect confidentiality, all patient information was de-identified and handled securely. The authors of this paper declare no conflicts of interest.

## RESULTS

### Description of Algorithms

We describe five distinct algorithms (three for cases; and two for controls): 1) code-based, imaging-confirmed cases, 2) code-based imaging-not-required cases, 3) NLP-based imaging-not-required cases, 4) code-based imaging-confirmed controls, and 5) code and NLP-based controls that did not require imaging. An overview of the inclusion and exclusion criteria for each of these algorithms is listed in Table 2 and details are described in Supplemental Tables 1-5. A full list of the CPT and ICD codes are listed in Supplemental Table 6.

**Table 2.**
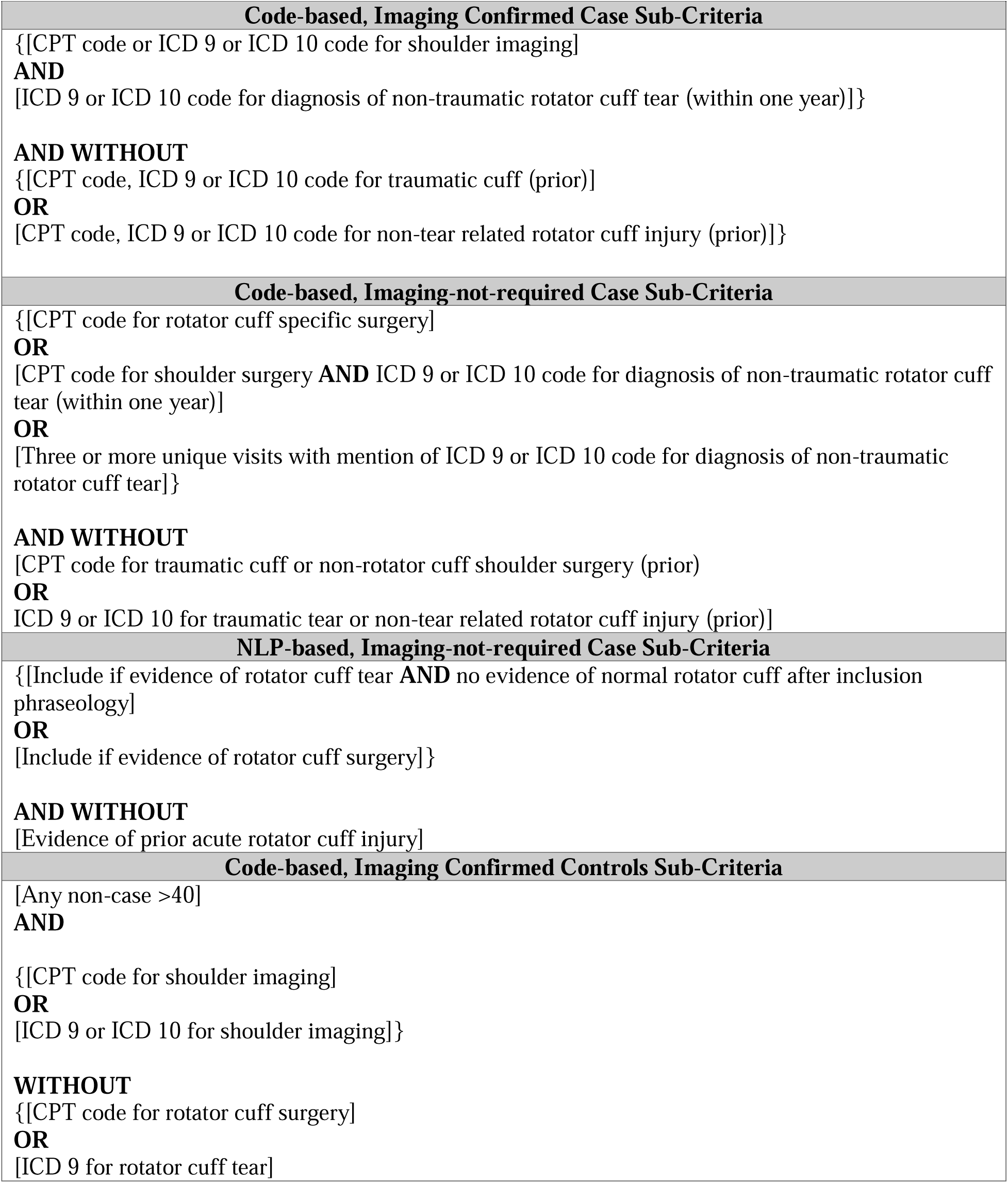
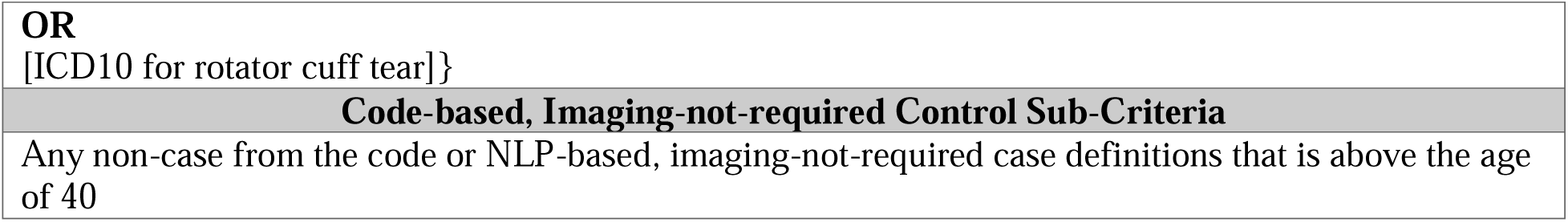
Algorithm Definitions.

#### Code-based, Imaging Confirmed Case Phenotype

The code-based imaging confirmed algorithm consists of one sub-criterion which was defined as the presence of a code (either ICD or CPT) for an imaging modality (MRI, CT-arthrogram, or ultrasound of the shoulder), followed by a code for rotator cuff tear within one year after the imaging code. This sub-criterion performed very well on the initial review and achieved a PPV of 92%. Because the algorithm was already performing well on the initial review, very little modification was made to the imaging confirmed case algorithm during an iterative review. The full imaging confirmed code case phenotype is listed in Supplemental Table 1.

#### Code-Based, Imaging-Not-Required, Case Phenotypes

The initial imaging-not-required code only case phenotype consisted of eight different sub-criteria (Supplemental Table 2) designed to comprehensively capture all methods that providers might use to document rotator cuff tear status in the electronic health record system. Possible approaches for the identification of cases in the initial algorithm consisted of 1) surgical confirmation, 2) provider diagnosis, and 3) physical therapy for rotator cuff tear. The eight different initial sub-criteria range from CPT codes specific to rotator cuff surgery (the most specific sub-criteria) to a single mention of an ICD code for a rotator cuff tear (the least specific sub-criteria). These initial algorithms were designed to capture as many cases of rotator cuff tear as possible and thus the predictive value of these algorithms after the first round of manual review varied greatly, from 31% PPV for the single ICD based sub-criteria to 89% PPV for the rotator cuff tear specific surgery CPT code-based sub-criteria (Supplemental Table 2).

After iterative review, four of the sub-criteria failed to meet the 85% PPV threshold and were excluded from consideration under final validation. These criteria included: both sub-criteria using codes for physical therapy (ICD based or CPT based) as a diagnostic modality for capturing cases (PPV 42% and 63% respectively) and the ICD-based definitions that only required one or two unique visits for rotator cuff tear only (PPV 31% and 36% respectively). Therefore, the final code-based imaging-not-required case algorithm consisted of four sub-criteria which are fully elucidated in Supplemental Table 2 and summarized in Table 2.

#### NLP-Based, Imaging-Not-Required Case Phenotypes

The initial imaging-not-required NLP case phenotype consisted of 15 different inclusion sub-criteria designed to capture the various ways that a medical professional might document the presence of a rotator cuff tear in the EHR. In addition to these 15 different affirmatory sub-criteria, the NLP algorithm also included seven different exclusion sub-criteria, designed to capture the various ways that a provider might document either a traumatic rotator cuff tear or the lack of a rotator cuff tear in the EHR. Unlike the initial code-based algorithms, which demonstrated a wide range of predictive values, the NLP-based algorithms had a much narrower span, ranging from 80% to 100%.

Only one sub-criterion was completely removed from the NLP-based code definition, the physical exam-based inclusion criterion, because it only captured 15 patients. However, because the remaining NLP-based algorithm performed remarkably well during initial review, it was modified very little from the original algorithm and only slight modifications to remove redundancy, include more specific terms and allow for plurality or past tense (such as supraspinatus AND teres minor were torn) were made. During this modification process two of the sub-criteria were condensed into one, ultimately resulting in a final NLP case algorithm of 13 inclusion terms and seven exclusion terms which are fully listed in Supplemental Table 3.

#### Code-based, Imaging-Confirmed Control Phenotypes

Similarly to the code-based imaging confirmed case algorithm, the code-based imaging confirmed control algorithm consisted only of one sub-criteria which was defined as any individual over the age of 40 who was not included as a code-based image-confirmed or imaging-not-required case, who additionally had an ICD or CPT code for imaging but who did NOT have the presence of an ICD code for a rotator cuff tear at any point after the imaging code. This sub-criterion consistently achieved 100% NPV on initial review and thus did not need to be modified during the iterative review process. The full imaging confirmed code control algorithm is listed in Supplemental Table 4.

#### Code + NLP-based, Imaging-Not-Required Control Phenotypes

Within the code-based, imaging-not-required control phenotype, the initial algorithm consisted of two different sub-criteria designed to capture the various levels of confidence in control status. The first sub-criteria included any non-case from the code or NLP non-image algorithms that were over the age of 40. The second sub-criteria consisted of individuals over 40 without evidence of a broader list of ICD/CPT codes for rotator cuff tear (Supplemental Tables 5 and 6). Both sub-criteria performed very well and had 95% and 90% NPV upon initial validation, respectively (Supplemental Table 5).

Though both sub-criteria of the code-based, imaging-not-required control definition performed well on iterative review and achieved greater than 85% predictive value, it was found that all controls captured by the second definition were also included in the first control definition. Thus, the second sub-criteria was dropped from the final algorithm due to redundancy. Therefore, the final code-based imaging-not-required control algorithm consisted of only one sub-criteria (any participant over the age of 40 who was not identified as a case by either the NLP or code-based non-image phenotypes).

### Final Algorithm Performances

The final algorithms underwent blinded review of 700 charts (400 cases and 300 controls). Among the 400 cases, 100 cases were selected from the code image-required algorithm, 100 were chosen from the NLP non-image required algorithm and 200 cases were selected from the code non-image required algorithm to allow sufficient cases to evaluate predictive values for each sub-criteria of the algorithm. Among the 300 controls, 100 controls were selected from the image-required algorithm and 200 were selected from the non-image required algorithm. Three records (one case and two controls) were excluded because they had no data available in the synthetic derivative and three additional records (all classified by the algorithm as controls) were deemed inconclusive because the data available in the chart was not adequate to rule in or out a rotator cuff tear. There was a 93.1% agreement among the reviewers with only 42 discordant charts.

Overall, the predictive value of the algorithm that did require imaging was 89.5% (179/200) with a positive predictive value of 89% for code-based imaging confirmed cases, and a negative predictive value of 90% for code-based imaging-not-required controls. For the imaging confirmed cases, the C-statistic was 0.921 (CI: 0.846-0.995). The imaging-not-required algorithm has an overall predictive value of 94.53% (464/494) with a positive predictive value of code-based imaging-not-required cases of 91.97%, NLP-based imaging-not-required cases of 89.00% and a negative predictive value of code and NLP-based imaging-not-required-controls of 100%. (Table 3) For the imaging-not-required algorithm, the C-statistic was 0.975 (CI: 0.946-1.005) and 0.945 (CI: 0.887-1.003) for code-based and NLP-based, respectively.

**Table 3.** Final Algorithm Counts and Predictive Values in VUMC SD.

| **Table 3. Final Algorithm Counts and Predictive Values in VUMC SD** |  |  |  |  |  |
| --- | --- | --- | --- | --- | --- |
|  | Imaging Confirmed |  | Imaging-not-required |  |  |
|  | Cases<br>(Code) | Controls<br>(Code) | Cases<br>(Code) | Cases<br>(NLP) | Controls<br>(Code + NLP) |
| **Total Count (N)** | 2,632 | 13,163 | 8,482 | 54,846 | 1,756,873 |
| **Predictive Value %**<br>**(# correctly classified/ total**<br>**randomly selected from**<br>**algorithm classification)** | 89%<br>(89/100) | 90%<br>(90/100) | 92%<br>(183/199) | 89%<br>(89/100) | 100%<br>(195/195) |

### External Validation

When applied to the pre-selected cohort at UTSW, the algorithm identified 420 true positives (TP), 26 false negatives (FN), 11 false positives (FP), and 35 true negatives (TN). This resulted in a sensitivity of 94%, specificity of 76%, and an accuracy of 92%. Further details of the external validation process are described in subsequent studies.

## DISCUSSION

In this study, we present two robust phenotypic algorithms for the classification of DCT in EHR-based systems with an predictive values over 90%, that performed well in external validation. Accurate phenotypic definitions are of paramount importance in achieving reliable results, particularly in the context of data derived for research from resources such as electronic health care records that were not collected for administrative, billing, and clinical assessment. One of the major limitations of current rotator cuff research in the context of using EHR data for research is the lack of clearly defined algorithms that identify DCT cases and controls. Furthermore, to our knowledge no study has developed and compared advantages, pitfalls, and consequences of decision criteria such as the use of an imaging requirement, or NLP in determining the size and composition of the study populations. The development and implementation of the algorithms outlined above, represent a significant improvement over prior strategies, that are reliant on ICD and CPT codes alone as results of the initial validation demonstrated that reliance on single occurrence of ICD codes, as is the current standard, has extremely poor predictive values. In contrast, the addition of temporal and frequency requirements, and NLP in the final algorithms substantially increased predictive value of the algorithms. This marked enhancement in performance opens the door for EHR based studies of unprecedented size. It overcomes the current limitations of poor phenotypic specificity of conventional coding methodologies and the time constraints of manual chart review.

One of the key strengths of this study is the versatility of the algorithms, as they offer the choice of using various combinations of structured and unstructured data, making it adaptable across different EHRs. The optional image requirements and ability to include NLP, allow for flexibility in application, catering to the diverse data structures and levels of query across various EHRs. This flexibility not only enhances the algorithm’s generalizability but also contributes to its utility in a broad range of clinical contexts, addressing the inherent variability in electronic health record formats and information availability. Table 4 outlines each of the algorithms as well as their advantages and draw backs.

**Table 4.** Algorithm Criteria and A Priori Comparison.

| **Table 4. Algorithm Criteria and A Priori Comparison** |  |  |  |
| --- | --- | --- | --- |
| **Cases** |  |  |  |
|  | **Imaging-not-required** |  | **Imaging Confirmed** |
|  | **Code Only** | **NLP** | **Code Only** |
| **Criteria** | ICD 9/10+CPT codes for rotator cuff tear | Language in the chart noting the presence of rotator cuff tear | ICD 9/10+CPT codes for imaging followed by codes for rotator cuff tear within one year after imaging code |
| **Advantages** | • More broadly applicable to EHRs | • Larger sample size | • Less misclassification |
| **Disadvantages** | • Higher potential for misclassification | • Fewer EHRs have NLP capabilities | • Smaller sample size<br>• Few EHRs have image querying abilities |
| **Controls** |  |  |  |
|  | **Imaging-not-required** |  | **Imaging Confirmed** |
|  | **Code + NLP** |  | **Code Only** |
| **Criteria** | Lack of ICD 9/10+CPT codes for rotator cuff tear and does not meet either CODE or NLP non-image required case definition |  | Lack of ICD 9/10+CPT Codes for rotator cuff tear and ICD/CPT for imaging without code for cuff tear |
| **Advantages** | • Larger sample size;<br>• More broadly applicable to EHRs<br>• Less risk of selection bias |  | • Less risk of misclassification |
| **Disadvantages** | • Higher risk of misclassification |  | • Limited sample size;<br>• Few EHRs have image querying abilities<br>• Higher risk of selection bias |

For EHR systems with the capability to include unstructured data, in the form of NLP, the integration of NLP into the non-image required algorithm significantly expanded the sample size of cases without meaningful compromise in predictive value. This augmentation is particularly noteworthy in genetic research as these studies often require extremely large sample sizes to detect a modest magnitude of associations between genetic variants and outcomes of interest. Large sample sizes enable the identification of these subtle genetic influences, contributing to a more comprehensive understanding of the genetic basis of traits and diseases. Additionally, given that genetic effects may differ among subgroups within a population (e.g., different ethnic groups or age categories), larger sample sizes provide the opportunity to investigate these subgroups and identify potential genetic heterogeneity, leading to a more nuanced understanding of genetic influences on traits. With a diverse and large sample, researchers can draw more reliable conclusions about genetic associations that may be generalizable to diverse populations or highlight key differences that exist across genetic ancestries.

Importantly, however, a delicate balance often exists between achieving precision of phenotypic definition and sample size, wherein increasing the stringency of a phenotypic definition often translates to lower case counts. In the instance of the algorithms outlined above, since the gold standard for rotator cuff tear diagnosis is imaging, the image verification component of the algorithm was designed to increase confidence in assessing case status while understanding that this would likely result in decreased sample size. After the final review, as expected, the sample size was significantly limited by the image requirement. However, surprisingly, the requirement for imaging in the algorithm did not result in a higher predictive value than that of the imaging-not-required algorithms. One potential reason for this difference is the stage of disease captured by each algorithm’s definition. Cases that have imaging documentation in the EHR were likely in the diagnostic phase of rotator cuff tear assessment when captured. Because of this, the imaging confirmed case definition captures cases with a degree of uncertainty as to diagnostic status, resulting in more false positive cases. On the other hand, those captured through imaging-not-required algorithms have often been previously diagnosed and are being captured either through referral for surgical procedures or from documentation of historical tear/repair. It is impossible for a provider to note all components of a normal medical history/physical exam and thus it is very unlikely that an intact rotator cuff tear would be documented for a patient in the imaging-not-required group.

## LIMITATIONS

There are several important considerations of this study that are inherent to the diagnostic heterogeneity of rotator cuff diseases. Firstly, while the requirement for imaging increased confidence in case status, it resulted in a meaningful reduction in sample size without improvement predictive value compared to non-imaging algorithms. This reduction may limit the generalizability of the findings. Additionally, imaging-confirmed cases, captured during the diagnostic phase, often involve greater diagnostic uncertainty, potentially leading to a higher number of false positives. On the other hand, the imaging-not-required algorithms might capture cases with a bias towards those diagnosed through referrals or historical documentation, possibly skewing results.

Lastly, the differences in disease stages between the algorithms could introduce variability in predictive values. Namely, the proposed algorithms are designed to be applied to large EHR based studies where the target population is the general public. However, the CuffGEN study relies on patients who presented to an orthopedic clinic for shoulder pain and underwent shoulder imaging. Because of this, the controls captured in this study inherently had some form of shoulder pathology that would indicate imaging and likely not representative of the general population. Though this does present a limitation of the study, it results in bias towards underperformance of the algorithm and is likely driving the lower specificity observed in external validation.

## CONCLUSIONS

The development and implementation of the algorithms outlined above, represent a significant improvement over prior strategies, that are reliant on ICD and CPT codes alone. The initial validation demonstrated that reliance on single occurrences of ICD codes, as is the current standard, have extremely poor predictive values. All algorithms exceeded the predictive value thresholds and demonstrated satisfactory results when applied to an outside EHR system. This marked enhancement in performance opens the door for EHR based studies of unprecedented size. It overcomes the current limitations of poor phenotypic specificity of conventional coding methodologies and the time constraints of manual chart review. By developing a novel informatics framework, this project provided evidenced-based solutions that substantially improve the accuracy and scalability of patient phenotype identification, thereby facilitating more comprehensive and reliable large scale clinical research.

## Data Availability

All data produced in the present study are available upon reasonable request to the authors

## Contributions/Acknowledgements

All authors contributed meaningfully to either to data collection, study design, or analysis. All authors have revised the manuscript and approved the final document.

## Data Sharing

All data pertaining to this publication is available upon request and approval.

## Ethical Approval

This study was approved by the VUMC IRB (#140857 and #060109) and the UTSW IRB (#STU-2020-0572 and #STU-2020-0978).

## Patient Involvement

Retrospective and prospective data was collected for patients who agreed to participate and signed informed consent. There were no additional interventions or treatments offered to patients in this study.

## Supplemental Figures/Tables

**Supplemental Table 1.** Code Image-Based Case Sub-Criteria.

Supplemental Figures/Tables:
| **Supplemental Table 1. Code Image-Based Case Sub-Criteria** |  |  |
| --- | --- | --- |
| Sub-Criteria Definition |  | Initial Validation Predictive Value |
| **1** | **CPT code for Shoulder imaging**<br>**OR**<br>**ICD 9 or 10 for Shoulder imaging**<br>**and**<br>**ICD 9 or 10 Code for Diagnosis of Non-Traumatic Rotator Cuff Tear (Within one year)**<br>**without**<br>**CPT Code for Traumatic Cuff or Non-Rotator Cuff Shoulder Surgery (PRIOR)**<br>**without**<br>**ICD 9 or 10 for Traumatic Tear or Non-Tear Related Rotator Cuff Injury (PRIOR)** | **92%** |
| This version of the algorithm underwent several iterative validations and the predictive values listed here represent the highest predictive value achieved by this sub-criterion before final validation. This sub-criterion is listed in bold because it achieved greater than the 85% predictive value threshold and was thus included in the final algorithm. |  |  |

**Supplemental Table 2.** Code-based Non-Image-Based Case Sub-Criteria.

| **Supplemental Table 2. Code-based Non-Image-Based Case Sub-Criteria** |  |  |
| --- | --- | --- |
| Sub-Criteria Definition |  | Initial Validation Predictive Value |
| **1** | **CPT Code for Rotator Cuff Specific Surgery**<br>**without**<br>**CPT Code for Traumatic Cuff or Non-Rotator Cuff Shoulder Surgery (PRIOR)**<br>**without**<br>**ICD 9 or 10 for Traumatic Tear or Non-Tear Related Rotator Cuff Injury (PRIOR)** | **95%** |
| **2** | **CPT code for Shoulder Surgery**<br>**and**<br>**ICD 9 or 10 Code for Diagnosis of Non-Traumatic Rotator Cuff Tear (Within one year)**<br>**without**<br>**CPT Code for Traumatic Cuff or Non-Rotator Cuff Shoulder Surgery (PRIOR)**<br>**without**<br>**ICD 9 or 10 for Traumatic Tear or Non-Tear Related Rotator Cuff Injury (PRIOR)** | **90%** |
| 3a | ICD 9 or 10 for Physical Therapy<br>and<br>ICD 9 or 10 Code for Diagnosis of Non-Traumatic Rotator Cuff Tear (Within one year)<br>without<br>CPT Code for Traumatic Cuff or Non-Rotator Cuff Shoulder Surgery (PRIOR)<br>without<br>ICD 9 or 10 for Traumatic Tear or Non-Tear Related Rotator Cuff Injury (PRIOR) | 42% |
| 3b | CPT code for Physical Therapy<br>and<br>ICD 9 or 10 Code for Diagnosis of Non-Traumatic Rotator Cuff Tear (Within one year)<br>without<br>CPT Code for Traumatic Cuff or Non-Rotator Cuff Shoulder Surgery (PRIOR)<br>without | 63% |
|  | ICD 9 or 10 for Traumatic Tear or Non-Tear Related Rotator Cuff Injury (PRIOR) |  |
| 4a | One unique visit with mention of ICD 9 or 10 Code for Diagnosis of Non-Traumatic Rotator Cuff Tear (Within one year)<br>without<br>CPT Code for Traumatic Cuff or Non-Rotator Cuff Shoulder Surgery (PRIOR)<br>without<br>ICD 9 or 10 for Traumatic Tear or Non-Tear Related Rotator Cuff Injury (PRIOR) | 31% |
| 4b | Two unique visits with mention of ICD 9 or 10 Code for Diagnosis of Non-Traumatic Rotator Cuff Tear (Within one year)<br>without<br>CPT Code for Traumatic Cuff or Non-Rotator Cuff Shoulder Surgery (PRIOR)<br>without<br>ICD 9 or 10 for Traumatic Tear or Non-Tear Related Rotator Cuff Injury (PRIOR) | 36% |
| 4c | **Three unique visits with mention of ICD 9 or 10 Code for Diagnosis of Non-Traumatic Rotator Cuff Tear (Within one year)**<br>**without**<br>**CPT Code for Traumatic Cuff or Non-Rotator Cuff Shoulder Surgery (PRIOR)**<br>**without**<br>**ICD 9 or 10 for Traumatic Tear or Non-Tear Related Rotator Cuff Injury (PRIOR)** | **87%** |
| 4d | **Four or more unique visits with mention of ICD 9 or 10 Code for Diagnosis of Non-Traumatic Rotator Cuff Tear (Within one year)**<br>**without**<br>**CPT Code for Traumatic Cuff or Non-Rotator Cuff Shoulder Surgery (PRIOR)**<br>**without**<br>**ICD 9 or 10 for Traumatic Tear or Non-Tear Related Rotator Cuff Injury (PRIOR)** | **92%** |
| This version of the algorithm underwent several iterative validations and the predictive values listed here represent the highest predictive value achieved by each sub-criteria before final validation. The sub-criteria listed in in bold are those that met the initial predictive value threshold of 85% and were included in the final algorithm. |  |  |

**Supplemental Table 3.** NLP-based Image Not Required Case Definition.

| Supplemental Table 3. NLP-based Image Not Required Case Definition |  |  |
| --- | --- | --- |
| Sub-Criteria Definition |  | Initial Validation Predictive Values |
| Exclusion Criteria* |  |  |
| 1 | Rotator cuff (is)... normal |  |
| 2 | Normal rotator cuff |  |
| 3 | No (evidence of/discernable/obvious/significant/ partial/ full-thickness/) rotator cuff tear/avulsion/pathology/abnormality(s) |  |
| 4 | Rotator cuff ... intact |  |
| 5 | Traumatic tear of (right/left) rotator cuff |  |
| 6 | No tear/torn (of) rotator cuff |  |
| 7 | Rotator cuff... no tear |  |
| Inclusion Criteria |  |  |
| 1 | (complete/partial/full/entire/incomplete/mild/massive/bilateral/small) (thickness) rotator cuff/cuff/supraspinatus (and)/infraspinatus (and)/subscapularis (and)/teres minor(and) (partial/partially/completely/complete) (tendon (s)) (is/has been/has a)(partial/partially/full/fully/complete/completely) (thickness) | 100% |
|  | **torn/tear/avulsion/avulsed** |  |
| **2** | **(complete/partial/full/entire/incomplete/mild/massive/bilateral/small) (thickness) tear of the (tendon(s) of the) rotator cuff/ cuff/Supraspinatus (and)/infraspinatus (and)/subscapularis (and)/teres minor (and)** | **90%** |
| **3** | **(completely/massively/entirely/fully/ partially) torn rotator cuff/cuff/supraspinatus (and)/infraspinatus (and)/subscapularis (and)/teres minor (and)** | **90%** |
| **4** | **(complete/massive/entire/full/partial) defect of (the) rotator cuff/cuff/supraspinatus (and) (and)/infraspinatus(and)/subscapularis (and)/teres minor (and)** | **100%** |
| **5** | **(complete/massive/entire/full/ partial) rotator cuff/cuff/supraspinatus(and)/infraspinatus(and)/subscapularis(and)/teres minor(and) defect(s)** | **100%** |
| 6** | (full/ partial) rotator cuff/cuff/supraspinatus/infraspinatus/subscapularis/teres minor (tear) repair | 100% |
| **7** | **rotator cuff/cuff/supraspinatus (and)/infraspinatus (and)/subscapularis (and)/teres minor (and)repair(ed) (arthroscopically)** | **100%** |
| **8** | **(arthroscopically) repair(ed) (of)(the) rotator cuff/cuff/supraspinatus (and)/infraspinatus (and)/subscapularis (and)/teres minor (and)** | **100%** |
| **9** | **arthroplasty of rotator cuff/cuff/supraspinatus (and)/infraspinatus (and)/subscapularis (and)/teres minor (and)** | **100%** |
| **10** | **rotator cuff/cuff/supraspinatus (and)/infraspinatus (and)/subscapularis (and)/teres minor (and) arthroplasty** | **100%** |
| **11\*\*** | **(surgery) (of the) arthroplastic/arthroplasty/arthroscopic (surgery) (of the)? rotator cuff/cuff/supraspinatus (and)/infraspinatus (and)/subscapularis (and)/teres minor (and) ... repair(ed)** | **100%** |
| **12** | **rotator cuff/cuff/supraspinatus (and)/infraspinatus (and)/subscapularis (and)/teres minor (and) surgery** | **100%** |
| **13** | **(surgical/surgically) (repair(ed)) (of) (the) rotator cuff/cuff/supraspinatus (and)/infraspinatus (and)/subscapularis (and)/teres minor(and) tear (arthroscopic/arthroplastic) repair/arthroplasty** | **100%** |
| **14** | **shoulder arthroplasty with repair of (the) rotator cuff/cuff/supraspinatus (and)/infraspinatus (and)/subscapularis (and)/teres minor (and) –** | **100%** |
| 15 | Physical examination:... Rotator cuff/Empty Can/ Neer/Hawkins/ Lift off/ Belly Press/ Drop Arm/ Bear Hug/ ... positive/+ | 80% |
| This version of the algorithm underwent several iterative validations and the predictive values listed here represent the highest predictive value achieved by each sub-criteria before final validation. The sub-criteria listed in bold met the initial predictive value threshold of 85% and were included in the final algorithm.<br>() = optional terms<br>/ = interchangeable terms<br>... = allow for space between terms – 20 characters for exclusion criteria and 55 characters for inclusion criteria<br>*Exclusion terms were applied to all sub-criteria before inclusion criteria<br>** Sub-criterion 6 was removed because it was captured entirely by sub criterion 11 |  |  |

**Supplemental Table 4.** Code-based Image Confirmed Control Sub-Criteria.

| **Supplemental Table 4. Code-based Image Confirmed Control Sub-Criteria** |  |  |
| --- | --- | --- |
| Sub-Criteria Definition |  | Initial Validation Predictive Value |
| **3** | **Any non-case >40**<br>**With** | **100%** |
|  | CPT code for Shoulder imaging<br>OR<br>ICD 9 or 10 for Shoulder imaging<br>Without<br>CPT:<br>i. CPT code 23412: repair of ruptured musculotendinous cuff (eg. Rotator cuff) open; chronic<br>ii. OR CPT code 23420: reconstruction of complete shoulder (rotator) cuff avulsion; chronic<br>iii. CPT code 23410: repair of ruptured musculotendinous cuff (eg. Rotator cuff) open; acute<br>iv. CPT code 29827 and 29828: Arthroscopy shoulder, surgical; with rotator cuff repair<br>ICD 9<br>i. 727.61 Complete rupture of rotator cuff<br>ii. 726.13 Partial tear of rotator cuff<br>iii. 726.1 Rotator cuff syndrome of shoulder and allied disorders<br>ICD10<br>i. M75.120 Complete rotator cuff tear or rupture of unspecified shoulder, not specified as traumatic<br>ii. M75.121 Complete rotator cuff tear or rupture of right shoulder, not specified as traumatic<br>iii. M75.122 Complete rotator cuff tear or rupture of left shoulder, not specified as traumatic<br>iv. M75.110 Incomplete rotator cuff tear or rupture of unspecified shoulder, not specified as traumatic<br>v. M75.111 Incomplete rotator cuff tear or rupture of right shoulder, not specified as traumatic<br>vi. M75.112 Incomplete rotator cuff tear or rupture of left shoulder, not specified as traumatic<br>vii. M75.100 Unspecified rotator cuff tear or rupture of unspecified shoulder, not specified as traumatic<br>viii. M75.101 Unspecified rotator cuff tear or rupture of right shoulder, not specified as traumatic<br>**M75.102 Unspecified rotator cuff tear or rupture of left shoulder, not specified as traumatic** |  |
| This version of the algorithm underwent several iterative validations and the predictive values listed here represent the highest predictive value achieved by this sub-criterion before final validation. This sub-criterion is bolded because it met the 85% predictive value threshold and was included in the final algorithm. |  |  |

**Supplemental Table 5.** Code-based Image Not Required Control Sub-Criteria.

|  |  |  |
| --- | --- | --- |
| Sub-Criteria Definition |  | Initial Validation Predictive Value |
| **1** | **Any non-case from the Code or NLP non image required case definitions that is above the age of 40** | **95%** |
| 2 | Anyone above the age of 40, without:<br>CPT: | 90% |
|  | v. CPT code 23412: repair of ruptured musculotendinous cuff (eg. Rotator cuff) open; chronic<br>vi. OR CPT code 23420: reconstruction of complete shoulder (rotator) cuff avulsion; chronic<br>vii. CPT code 23410: repair of ruptured musculotendinous cuff (eg. Rotator cuff) open; acute<br>viii. CPT code 29827 and 29828: Arthroscopy shoulder, surgical; with rotator cuff repair<br>ICD 9<br>iv. 727.61 Complete rupture of rotator cuff<br>v. 726.13 Partial tear of rotator cuff<br>vi. 726.1 Rotator cuff syndrome of shoulder and allied disorders<br>ICD10<br>ix. M75.120 Complete rotator cuff tear or rupture of unspecified shoulder, not specified as traumatic<br>x. M75.121 Complete rotator cuff tear or rupture of right shoulder, not specified as traumatic<br>xi. M75.122 Complete rotator cuff tear or rupture of left shoulder, not specified as traumatic<br>xii. M75.110 Incomplete rotator cuff tear or rupture of unpsecified shoulder, not specified as traumatic<br>xiii. M75.111 Incomplete rotator cuff tear or rupture of right shoulder, not specified as traumatic<br>xiv. M75.112 Incomplete rotator cuff tear or rupture of left shoulder, not specified as traumatic<br>xv. M75.100 Unspecified rotator cuff tear or rupture of unspecified shoulder, not specified as traumatic<br>xvi. M75.101 Unspecified rotator cuff tear or rupture of right shoulder, not specified as traumatic<br>xvii. M75.102 Unspecified rotator cuff tear or rupture of left shoulder, not specified as traumatic |  |
| This version of the algorithm underwent several iterative validations and the predictive values listed here represent the highest predictive value achieved by these sub-criteria before final validation. Both sub-criterion achieved greater than the 85% predictive value threshold, but the first criterion was the only one included in the final algorithm because it was found to capture everyone listed in the second criterion. |  |  |

**Supplemental Table 6.** Full List of ICD and CPT Codes Used in Code-based Algorithms.

| Code Type | Code | Code Explanation |
| --- | --- | --- |
| **Codes for Rotator Cuff Tear** |  |  |
| CPT | 23412 | repair of ruptured musculotendinous cuff (eg. Rotator cuff) open; chronic |
| CPT | 23420 | reconstruction of complete shoulder (rotator) cuff avulsion; chronic |
| CPT | 23397 | Under Repair, Revision, and/or Reconstruction Procedures on the Shoulder |
| CPT | 29826 | Arthroscopy, shoulder, surgical; decompression of subacromial space with partial |
| CPT | 29901 | Under Endoscopy/Arthroscopy Procedures on the Musculoskeletal System |
| CPT | 29827 | Arthroscopy shoulder, surgical; with rotator cuff repair |
| CPT | 29828 | Arthroscopy shoulder, surgical; with rotator cuff repair |
| CPT | 29822 | Arthroscopy, shoulder, surgical; debridement, limited includes debridement of soft or hard tissue |
| ICD9CM | 727.61 | Complete rupture of rotator cuff |
| ICD9CM | 726.13 | Partial tear of rotator cuff |
| ICD9CM | 83.63 | Rotator cuff repair |
| ICD10CM | M75.120 | Complete rotator cuff tear or rupture of unspecified shoulder, not specified as traumatic |
| ICD10CM | M75.121 | Complete rotator cuff tear or rupture of right shoulder, not specified as traumatic |
| ICD10CM | M75.122 | Complete rotator cuff tear or rupture of left shoulder, not specified as traumatic |
| ICD10CM | M75.110 | Incomplete rotator cuff tear or rupture of unspecified shoulder, not specified as traumatic |
| ICD10CM | M75.111 | Incomplete rotator cuff tear or rupture of right shoulder, not specified as traumatic |
| ICD10CM | M75.112 | Incomplete rotator cuff tear or rupture of left shoulder, not specified as traumatic |
| ICD10CM | M75.100 | Unspecified rotator cuff tear or rupture of unspecified shoulder, not specified as traumatic |
| ICD10CM | M75.101 | Unspecified rotator cuff tear or rupture of right shoulder, not specified as traumatic |
| ICD10CM | M75.102 | Unspecified rotator cuff tear or rupture of left shoulder, not specified as traumatic |
| **Codes for Rotator Cuff Exclusions (Traumatic Tear)** |  |  |
| CPT | 23410 | repair of ruptured musculotendinous cuff (eg. Rotator cuff) open; acute |
| CPT | 24341 | repair, tendon or muscle, upper arm or elbow, each tendon or muscle, primary or secondary (excludes Rotator cuff) |
| ICD9CM | 840.3 | Infraspinatus (muscle) (tendon) sprain |
| ICD9CM | 840.4 | Rotator cuff (capsule) sprain |
| ICD9CM | 840.5 | Subscapularis (muscle) sprain |
| ICD9CM | 840.6 | Supraspinatus (muscle) (tendon) sprain |
| ICD10CM | S46.011A | Strain of muscle(s) and tendon(s) of the rotator cuff of right shoulder, initial encounter |
| ICD10CM | S46.011D | Strain of muscle(s) and tendon(s) of the rotator cuff of right shoulder, subsequent encounter |
| ICD10CM | S46.011S | Strain of muscle(s) and tendon(s) of the rotator cuff of right shoulder, sequela |
| ICD10CM | S46.012A | Strain of muscle(s) and tendon(s) of the rotator cuff of left shoulder, initial encounter |
| ICD10CM | S46.012D | Strain of muscle(s) and tendon(s) of the rotator cuff of left shoulder, sequential encounter |
| ICD10CM | S46.012S | Strain of muscle(s) and tendon(s) of the rotator cuff of left shoulder, sequela |
| ICD10CM | S46.011A | Strain of muscle(s) and tendon(s) of the rotator cuff of unspecified shoulder, initial encounter |
| ICD10CM | S46.011D | Strain of muscle(s) and tendon(s) of the rotator cuff of unspecified shoulder, subsequent encounter |
| ICD10CM | S46.011S | Strain of muscle(s) and tendon(s) of the rotator cuff of unspecified shoulder, sequela |
| ICD10CM | S46.021A | Laceration of muscle(s) and tendon(s) of the rotator cuff of the right shoulder, initial encounter |
| ICD10CM | S46.021D | Laceration of muscle(s) and tendon(s) of the rotator cuff of the right shoulder, Sequential encounter |
| ICD10CM | S46.021S | Laceration of muscle(s) and tendon(s) of the rotator cuff of the right shoulder, sequela |
| ICD10CM | S46.022A | Laceration of muscle(s) and tendon(s) of the rotator cuff of the left shoulder, initial encounter |
| ICD10CM | S46.022D | Laceration of muscle(s) and tendon(s) of the rotator cuff of the left shoulder, Sequential encounter |
| ICD10CM | S46.022S | Laceration of muscle(s) and tendon(s) of the rotator cuff of the left shoulder, sequela |
| ICD10CM | S46.029A | Laceration of muscle(s) and tendon(s) of the rotator cuff of unspecified shoulder, initial encounter |
| ICD10CM | S46.029D | Laceration of muscle(s) and tendon(s) of the rotator cuff of unspecified shoulder, Sequential encounter |
| ICD10CM | S46.029S | Laceration of muscle(s) and tendon(s) of the rotator cuff of unspecified shoulder, sequela |
| ICD10CM | S43.421A | Sprain of muscle(s) and tendon(s) of the rotator cuff of right shoulder, initial encounter |
| ICD10CM | S43.421D | Sprain of muscle(s) and tendon(s) of the rotator cuff of right shoulder, sequential encounter |
| ICD10CM | S43.421S | Sprain of muscle(s) and tendon(s) of the rotator cuff of right shoulder, sequela |
| ICD10CM | S43.422A | Sprain of muscle(s) and tendon(s) of the rotator cuff of left shoulder, initial encounter |
| ICD10CM | S43.422D | Sprain of muscle(s) and tendon(s) of the rotator cuff of left shoulder, sequential encounter |
| ICD10CM | S43.422S | Sprain of muscle(s) and tendon(s) of the rotator cuff of left shoulder, sequela |
| ICD10CM | S43.429A | Sprain of muscle(s) and tendon(s) of the rotator cuff of unspecified shoulder, initial encounter |
| ICD10CM | S43.429D | Sprain of muscle(s) and tendon(s) of the rotator cuff of unspecified shoulder, sequential encounter |
| ICD10CM | S43.429S | Sprain of muscle(s) and tendon(s) of the rotator cuff of unspecified shoulder, sequela |
| ICD10CM | M12.511 | Traumatic Arthropathy, right shoulder |
| ICD10CM | M12.512 | Traumatic Arthropathy, left shoulder |
| ICD10CM | M12.519 | Traumatic Arthropathy, unspecified shoulder |
| ICD10CM | M75.30 | Calcific Tendinitis of unspecified shoulder |
| ICD10CM | M75.31 | Calcific Tendinitis of right shoulder |
| ICD10CM | M75.32 | Calcific Tendinitis of left shoulder |
| Codes for Shoulder Imaging |  |  |
| CPT | 76880 | [Expired] Ultrasound, extremity, nonvascular, real time with image documentation |
| CPT | 78661 | Ultrasound, complete joint (ie. Joint space and peri-articular soft tissue structures) real time with image documentation |
| CPT | 78662 | Ultrasound, limited, joint or other non-vascular extremity structure (i.e. joint space, peri-articular tendon[s], muscle[s], nerve[s], other soft tissue structure[s], or soft tissue mass[es]) real time with image documentation |
| ICD9CM | 88.94 | Magnetic Resonance Imaging of Musculoskeletal |
| ICD9CM | 88.32 | Contrast arthrogram |
| ICD9CM | 88.7 | Diagnostic Ultrasound |
| ICD10CM | BP3EZZZ | MRI upper extremity left |
| ICD10CM | BP3FYZZ | MRI upper extremity left, with contrast |
| ICD10CM | BP3FY0Z | MRI upper extremity left, enhanced or unenhanced |
| ICD10CM | BP38ZZZ | MRI upper extremity right |
| ICD10CM | BP38YZZ | MRI upper extremity right, with contrast |
| ICD10CM | BP38Y0Z | MRI upper extremity right, enhanced or unenhanced |
| ICD10CM | BL33YZZZ | MRI upper extremity tendon |
| ICD10CM | BL33YZZ | MRI upper extremity tendon with contrast |
| ICD10CM | BL33Y0Z | MRI upper extremity tendon with contrast, enhanced or unenhanced |
| ICD10CM | BL30ZZZ | MRI upper extremity Connective tissue |
| ICD10CM | BL30YZZ | MRI upper extremity Connective tissue with contrast |
| ICD10CM | BL30Y0Z | MRI upper extremity Connective Tissue with contrast, enhanced or unenhanced |
| ICD10CM | B53NZZZ | MRI upper extremity vein |
| ICD10CM | B53NYZZ | MRI upper extremity vein with contrast |
| ICD10CM | B53NY0Z | MRI upper extremity vein with contrast enhanced or unenhanced |
| ICD10CM | B33KZZZ | MRI upper extremity artery |
| ICD10CM | B33KYZZ | MRI upper extremity artery with contrast |
| ICD10CM | B33KY0Z | MRI upper extremity artery with contrast enhanced or unenhanced |
| ICD10CM | B33JZZZ | MRI Left upper extremity artery |
| ICD10CM | B33JYZZ | MRI Left upper extremity artery with contrast |
| ICD10CM | B33JY0Z | MRI Left upper extremity artery with contrast enhanced or unenhanced |
| ICD10CM | B33HZZZ | MRI Right upper extremity artery |
| ICD10CM | B33HYZZ | MRI Right upper extremity artery with contrast |
| ICD10CM | B33HY0Z | MRI Right upper extremity artery with contrast enhanced or unenhanced |
| Codes for Non-Traumatic Tendon Injury |  |  |
| ICD9CM | 727.6 | Rupture of tendon nontraumatic |
| ICD9CM | 727.60 | Nontraumatic rupture of unspecified tendon |
| **Codes for Physical Therapy** |  |  |
| CPT | 90071 | [Expired] Physical therapy evaluation |
| CPT | 90072 | [Expired] Physical therapy re-evaluation |
| CPT | 97161 | Physical therapy, low complexity, typical time with patient and/or/family 10 mins |
| CPT | 97162 | Physical therapy, moderate complexity, typical time with patient and/or/family 30 mins |
| CPT | 97163 | Physical therapy, high complexity, typical time with patient and/or/family 45 mins |
| CPT | 97164 | Reevaluation of physical therapy established plan of care, requiring these components: An examination including a review of history and use of standardized tests and measures is required; and revised plan of care using a standardized patient assessment instrument or measurable assessment of functional outcome Typically 20 mins are spent with the patient or family. |
| ICD9CM | 93.0 | physical therapy, respiratory therapy, rehabilitation, and related procedures |
| ICD9CM | 93.00 | diagnostic physical therapy |
| ICD9CM | 93.09 | other diagnostic physical therapy procedure |
| ICD9CM | 93.1 | physical therapy procedure |
| ICD9CM | 93.2 | other physical therapy musculoskeletal procedures |
| ICD9CM | 93.38 | combined physical therapy without mention of components |
| ICD9CM | 93.39 | other physical therapy |
| ICD9CM | V57.1 | care involving other physical therapy |

## REFERENCES

1 Varacallo M, El Bitar Y, Mair SD. Rotator Cuff Syndrome. StatPearls. Treasure Island (FL): StatPearls Publishing 2023.

2 Karjalainen TV, Jain NB, Heikkinen J, et al. Surgery for rotator cuff tears. Cochrane Database Syst Rev. 2019;12:CD013502. doi: 10.1002/14651858.CD013502

3 Minagawa H, Yamamoto N, Abe H, et al. Prevalence of symptomatic and asymptomatic rotator cuff tears in the general population: From mass-screening in one village. Journal of Orthopaedics. 2013;10:8–12. doi: 10.1016/j.jor.2013.01.008

4 Tempelhof S, Rupp S, Seil R. Age-related prevalence of rotator cuff tears in asymptomatic shoulders. Journal of Shoulder and Elbow Surgery. 1999;8:296–9. doi: 10.1016/S1058-2746(99)90148-9

5 An H-J, Kim J-H, Yoon S, et al. Genome-Wide Association Study Identifies Genetic Variants Associated with Rotator Cuff Tear-A Pilot Study. Diagnostics (Basel). 2022;12:2497. doi: 10.3390/diagnostics12102497

6 Harvie P, Ostlere SJ, Teh J, et al. Genetic influences in the aetiology of tears of the rotator cuff. Sibling risk of a full-thickness tear. J Bone Joint Surg Br. 2004;86:696–700. doi: 10.1302/0301-620x.86b5.14747

7 Gwilym SE, Watkins B, Cooper CD, et al. Genetic influences in the progression of tears of the rotator cuff. J Bone Joint Surg Br. 2009;91:915–7. doi: 10.1302/0301-620X.91B7.22353

8 Tashjian R, Granger E, Hung M, et al. Incidence of familial tendon dysfunction in patients with full-thickness rotator cuff tears. OAJSM. 2014;137. doi: 10.2147/OAJSM.S63656

9 Motta G da R, Amaral MV, Rezende E, et al. Evidence of genetic variations associated with rotator cuff disease. J Shoulder Elbow Surg. 2014;23:227–35. doi: 10.1016/j.jse.2013.07.053

10 Teerlink CC, Cannon-Albright LA, Tashjian RZ. Significant association of full-thickness rotator cuff tears and estrogen-related receptor-β (ESRRB). J Shoulder Elbow Surg. 2015;24:e31–35. doi: 10.1016/j.jse.2014.06.052

11 Tashjian RZ, Granger EK, Farnham JM, et al. Genome-wide association study for rotator cuff tears identifies two significant single-nucleotide polymorphisms. Journal of Shoulder and Elbow Surgery. 2016;25:174–9. doi: 10.1016/j.jse.2015.07.005

12 Assunção JH, Godoy-Santos AL, Dos Santos MCLG, et al. Matrix Metalloproteases 1 and 3 Promoter Gene Polymorphism Is Associated With Rotator Cuff Tear. Clin Orthop Relat Res. 2017;475:1904–10. doi: 10.1007/s11999-017-5271-3

13 Kluger R, Burgstaller J, Vogl C, et al. Candidate gene approach identifies six SNPs in tenascin-C (TNC) associated with degenerative rotator cuff tears. J Orthop Res. 2017;35:894–901. doi: 10.1002/jor.23321

14 Longo UG, Margiotti K, Petrillo S, et al. Genetics of rotator cuff tears: no association of col5a1 gene in a case-control study. BMC Med Genet. 2018;19:217. doi: 10.1186/s12881-018-0727-1

15 Salles JI, Lopes LR, Duarte MEL, et al. Fc receptor-like 3 (-169T>C) polymorphism increases the risk of tendinopathy in volleyball athletes: a case control study. BMC Med Genet. 2018;19:119. doi: 10.1186/s12881-018-0633-6

16 Tashjian RZ, Kim SK, Roche MD, et al. Genetic variants associated with rotator cuff tearing utilizing multiple population-based genetic resources. J Shoulder Elbow Surg. 2021;30:520–31. doi: 10.1016/j.jse.2020.06.036

17 Tashjian RZ, Farnham JM, Albright FS, et al. Evidence for an inherited predisposition contributing to the risk for rotator cuff disease. J Bone Joint Surg Am. 2009;91:1136–42. doi: 10.2106/JBJS.H.00831

18 Roos TR, Roos AK, Avins AL, et al. Genome-wide association study identifies a locus associated with rotator cuff injury. PLoS One. 2017;12:e0189317. doi: 10.1371/journal.pone.0189317

19 Liu L, Yang F, Liao Y, et al. Exploration of causal relationship between shoulder impingement syndrome and rotator cuff injury: a bidirectional mendelian randomization. BMC Musculoskelet Disord. 2024;25:649. doi: 10.1186/s12891-024-07556-1

20 Ryan BL, Maddocks HL, McKay S, et al. Identifying musculoskeletal conditions in electronic medical records: a prevalence and validation study using the Deliver Primary Healthcare Information (DELPHI) database. BMC Musculoskelet Disord. 2019;20:187. doi: 10.1186/s12891-019-2568-2

21 Gao C, Fan R, Ayers GD, et al. Development and Validation of an Electronic Medical Record Algorithm to Identify Phenotypes of Rotator Cuff Tear. PM R. 2020;12:1099–105. doi: 10.1002/pmrj.12367

22 Yanik EL, Keener JD, Lin SJ, et al. Identification of a Novel Genetic Marker for Risk of Degenerative Rotator Cuff Disease Surgery in the UK Biobank. JBJS. 2021;103:1259–67. doi: 10.2106/JBJS.20.01474

23 Tashjian RZ, Granger EK, Farnham JM, et al. Genome-wide association study for rotator cuff tears identifies two significant single-nucleotide polymorphisms. J Shoulder Elbow Surg. 2016;25:174–9. doi: 10.1016/j.jse.2015.07.005

24 Gaziano JM, Concato J, Brophy M, et al. Million Veteran Program: A mega-biobank to study genetic influences on health and disease. Journal of Clinical Epidemiology. 2016;70:214–23. doi: 10.1016/j.jclinepi.2015.09.016

25 Sudlow C, Gallacher J, Allen N, et al. UK Biobank: An Open Access Resource for Identifying the Causes of a Wide Range of Complex Diseases of Middle and Old Age. PLoS Med. 2015;12:e1001779. doi: 10.1371/journal.pmed.1001779

26 Roden DM, Pulley JM, Basford MA, et al. Development of a large-scale de-identified DNA biobank to enable personalized medicine. Clin Pharmacol Ther. 2008;84:362–9. doi: 10.1038/clpt.2008.89

27 Breeyear JH, Mitchell SL, Nealon CL, et al. Development of electronic health record based algorithms to identify individuals with diabetic retinopathy. Journal of the American Medical Informatics Association. 2024;ocae213. doi: 10.1093/jamia/ocae213

28 Harrell FE, Califf RM, Pryor DB, et al. Evaluating the yield of medical tests. JAMA. 1982;247:2543– 6.

